# Fractality of tics as a quantitative assessment tool for Tourette syndrome

**DOI:** 10.1101/2021.07.29.21261345

**Authors:** Payton Beeler, Nicholas O. Jensen, Soyoung Kim, Amy Robichaux-Viehoever, Bradley L. Schlaggar, Deanna J. Greene, Kevin J. Black, Rajan Chakrabarty

**Author notes:** Corresponding authors: Rajan K. Chakrabarty and Kevin J. Black., **Email:**. **Competing Interest Statement:** The authors declare that they have no known competing financial interests or personal relationships that could have appeared to influence the work reported in this paper.

## Abstract

Tics manifest as brief, purposeless, and unintentional movements or noises that, for many individuals, can be suppressed temporarily with effort. Peterson and Leckman (1998) hypothesized that the chaotic temporal nature of tics could possess an inherent fractality, that is, have neighbor-to-neighbor correlation at all levels of time scale. However, demonstrating this phenomenon has eluded researchers for more than two decades, primarily because of the challenges associated with estimating the scale-invariant, power law exponent – called the fractal dimension D_f_ – from fractional Brownian noise. Here, we confirm this hypothesis and establish the fractality of tics by examining two tic time series datasets collected 6-12 months apart in children with tics, using one-dimensional random walk models. We find that D_f_ increases from ∼1.4 to 1.75 in order of decreasing tic severity, and is correlated with tic severity as measured by the YGTTS total tic score. We demonstrate D_f_ to be a sensitive parameter in examining the effect of several tic suppression conditions on the tic time series. We confirm the fractal nature of tics in Tourette syndrome (TS) and extend the finding to Provisional Tic Disorder. Our findings pave the way for utilizing the fractal nature of tics as a robust quantitative tool for estimating tic severity and treatment effectiveness, as well as a marker for differentiating typical from functional tics.

## Introduction

Tics are brief, purposeless, unintentional behaviors appearing as repeated movements of skeletal or vocal musculature, affecting more than 20% of all children (1, 2). Approximately 0.5% of children have Tourette syndrome (TS), which is diagnosed when both motor and vocal tics occur over a period of a year or longer (3). Tic disorders, including TS, are moderately heritable, but despite decades of active scientific research, no consensus has been reached on their pathophysiological foundations (4, 5). In 1998, Peterson and Leckman noted that tics tend to arise in clusters (bouts of several tic occurrences within a few seconds, separated by longer tic-free intervals), but also that at longer time scales, bouts of tics lasting several seconds similarly recur in grouped episodes over the courses of hours (6, 7). Such recursive behavior in turn extends to longer timespans (days, weeks, and months), maintaining self-similarity (Figure 1, left). Their observation suggests that the occurrence of tics could have an associated fractal pattern. Consequently, one would expect the occurrences to exhibit neighbor-to-neighbor correlation and follow a power curve with a scale-invariant exponent, the fractal dimension (D_f_). We are unaware of any previous attempts at estimating D_f_ of tics, which would not only provide quantitative insights into the chaotic nature of tic disorders, but also facilitate provide new ways for assessing the disorder quantitatively.

**Figure 1.**
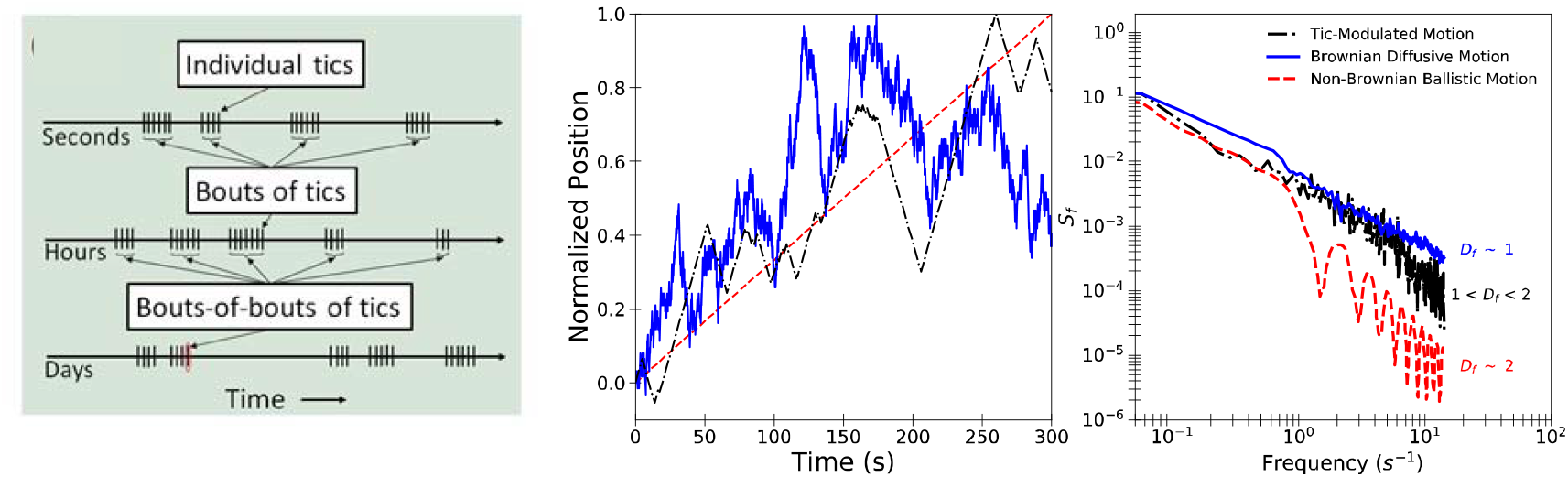
**(left)** Fractal pattern in the occurrence of tics, adapted from (7). **(center)** Trajectory of a tic-modulated random walk (black) is compared with that of Brownian diffusive motion (blue) and non-Brownian ballistic motion (red). The position of each walker at time *t* is normalized by the maximum displacement of the walker from *t* = 0 to *t* = 300, rendering the trajectories in one-dimensional space as a function of time. **(right)** Estimation of D_f_ (1<D_f_<2) for a walker undergoing diffusive motion (blue), non-Brownian motion (red), and tic-modulated motion (black).

We first recorded the timing of tics in patients during 5-minute video sessions under 4 conditions: free to tic (“baseline”), verbal request not to tic (“verbal”), immediate token rewards for 10-s tic-free periods (differential reinforcement of other, “DRO”), or tokens given at the same timing as in a previous DRO session regardless of current tic appearance (non-contingent reinforcement, “NCR”). Video sessions were conducted within the first 6 months after onset of tics (screening visit), when Provisional Tic Disorder can be diagnosed, and again at the 12 month anniversary of the first tic (12 month visit), when a chronic tic disorder (Tourette syndrome or Persistent Tic Disorder) can first be diagnosed. Next, we used a one-dimensional random walk model in which the movement direction of walkers is reversed with each tic exhibited by the patient, generating what is hereafter referred to as a “tic-modulated random walk”. The random walk model produced trajectories corresponding to each patient, visit, and condition. Further details regarding the random walk model appear in section S1 of the supporting information.

## Results

The center panel of Figure 1 shows the trajectory of a one-dimensional tic-modulated random walker with data obtained from one child with TS (black line). We then compared the trajectory of the tic-modulated random walker with two cases that represent the two extremes of chaotic timing. The first case is Brownian diffusive motion. Brownian diffusive motion represents the most chaotic case for a tic time series, in which at any time increment, the walker is equally likely to move in either direction. If a patient generates a tic time series that closely resembles Brownian diffusive motion, the patient is equally likely to tic or not tic at any given time. The second case is that of non-Brownian ballistic motion. Non-Brownian ballistic motion represents the most ordered case for a tic time series, where at any given time increment, the walker does not change directions and continues on its original path (i.e., the patient has a 0% chance of exhibiting a tic at any given time). When compared to the trajectory of a walker undergoing Brownian diffusion, the tic-modulated random walks appear to possess many scales. Unlike the Brownian diffusion case, some excursive episodes of the walker (*i*.*e*., longer periods without a tic) can be observed in the tic-modulated trajectories. Qualitatively speaking, the mobility of the tic-driven walker is stronger than that of Brownian diffusive motion but weaker than that of non-Brownian ballistic motion.

The fractal dimension of the tic time series was determined by analyzing the squared Fourier transform of the density autocorrelation function of the random walkers (S_f_) (8), which is given by:

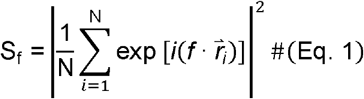

where 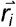 is the position of the *i*^th^ walker in *t*-*x* space, N is the number of time steps, and *f* is the frequency (8). Equation 1 is evaluated for each iteration of the random walk model. Specifically, S_f_ was found by first rotating the trajectory of the walker by an angle (*θ*), then solving equation 1 for a given frequency. The result of equation 1 is then averaged over 180 values of *θ* (evenly distributed between 0 and 2π), to give S_f_ as a function of frequency. The slope of S_f_ vs. *f* in log-log space then gives D_f_ of the tic time series (8). Figure 1 (right) shows examples of S_q_ for a tic-driven walker, Brownian diffusion, and non-Brownian ballistic motion. Non-Brownian ballistic motion and Brownian diffusion can be respectively quantified by D_f_ = 2 and D_f_ = 1. Simulated random walks define an expected range for D_f_ that correlates with tic severity, and experimentally measured tic time series can be reliably interpreted within this domain.

Figure 2a shows that for individual patients, the change in D_f_ of the tic time series is correlated with change in tic severity, as measured by the Yale Global Tic Severity Scale total tic score (TTS) between screening and 12 month visits (*r* = -0.33, *p* = 0.03). Patients with decreased TTS between screening and 12 month visits (greater improvement in tics) also had increased D_f_ of the tic time series. Given these results, D_f_ of the tic time series can be used to measure tic severity, with more severe tics having D_f_ ≈ 1, and less severe tics having D_f_ ≈ 2. Figure 2b demonstrates the overall effectiveness of various tic suppression methods using the average D_f_ of the tic-modulated walkers. Figure 2a shows that during screening visits, DRO and verbal conditions were effective suppression techniques, marked by a statistically significant increase in D_f_ (*p*’s < 0.01). NCR’s effects on D_f_ were not statistically significant. Twelve months after the onset of tics, DRO and verbal suppression conditions led to statistically significant increases in D_f_ (*p*’s < 0.01). Reproducibility of D_f_ and repeatability of conclusions were confirmed by co-author ARV (Figure 2 c-d), who independently recorded tic timing from the video recordings blind to visit or video condition (9). D_f_ from the two independent raters was highly correlated (*r* = 0.71, *N* = 194 sessions from screening visits).

**Figure 2.**
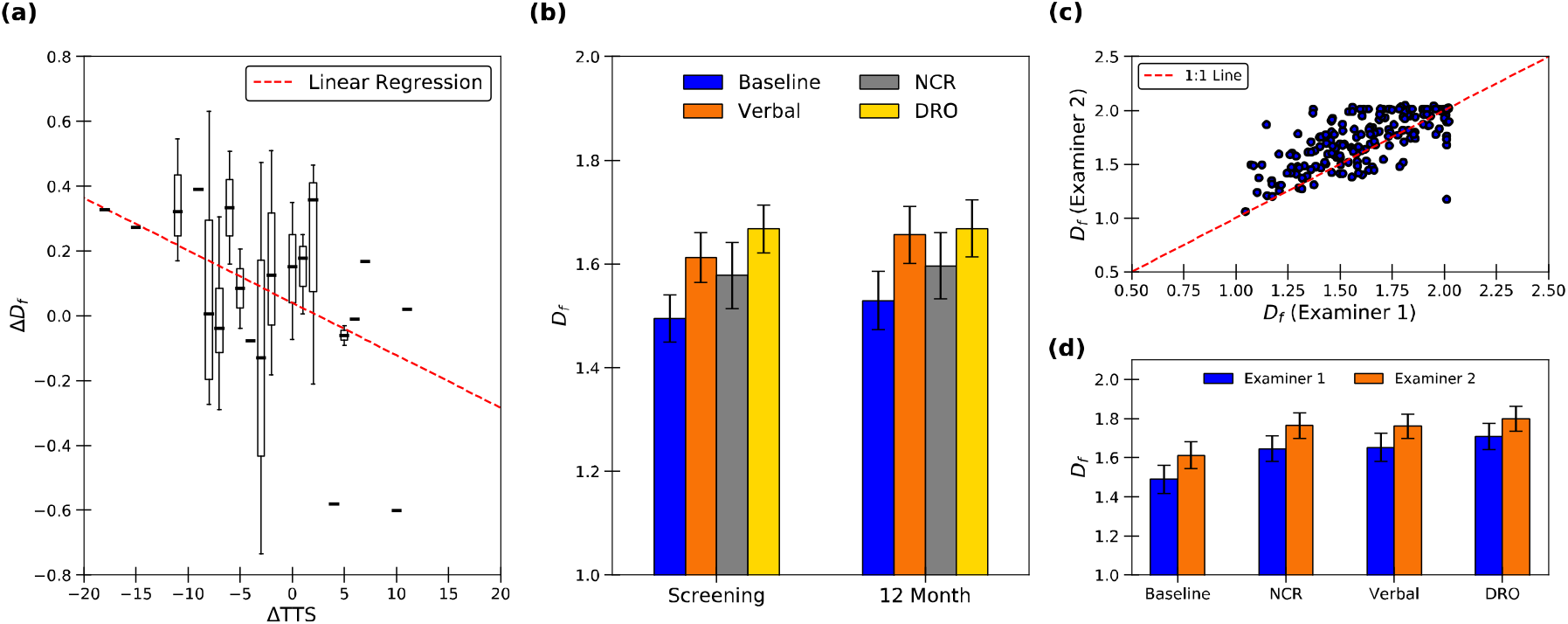
**(a)** Change in fractal dimension (D_f_) between screening and 12 month visit as a function of change in YGTSS total tic score (TTS). The red dashed line shows linear regression of ΔD_f_ vs ΔTTS, *r* = -0.333 **(b)** Comparison of average D_f_ for the patients under various suppression conditions during screening visits shows that DRO led to the most effective tic suppression (largest D_f_). Additionally, the fractal dimension of all suppression conditions increased at 12 months (when most patients met diagnostic criteria for TS). **(c-d)** Double-blind reproducibility of D_f_ as an assessment tool. Comparison of D_f_ for tic time series generated by 2 examiners, with examiner 2 being blind to visit and condition. Good agreement is observed between examiners, verifying inter-rater reliability of results. All error bars show 95% confidence intervals.

## Discussion

Here we validate the hypothesis of fractal timing of tics in TS first reported over 20 years ago (6), and extend that observation for the first time to tics shortly after they appear (Provisional Tic Disorder). We first show that D_f_ of the tic time series is correlated with a standard clinical measure of tic severity (TTS), suggesting that D_f_ of the tic time series can be used as an objective measure of tic severity. Additionally, we measure for the first time the effect of tic suppression on the temporal dynamics of tic occurrence by quantification of D_f_. Using this analytical framework, we further demonstrate that various tic suppression techniques have an effect on this relationship, with increased effectiveness reflected by increased D_f_. Overall, DRO and verbal instruction were effective tic suppression conditions during screening and 12 month visits, with DRO being the most effective. In addition, all conditions showed increased D_f_ at 12 month visits compared to screening visits, which may be attributed to the passage of time, with continuing cognitive development and additional practice with environmental tic suppression resulting in improved tic inhibition in the social environment. Finally, we demonstrate the robustness of this parameter via the congruence with our results by a double-blind rater. Future work should be directed to extend the scaling analysis to longer timespans, since qualitatively the temporal dynamics of tics have been observed to maintain self-similarity over months and even years. This method for analyzing the timing of tic occurrence shows promise for documenting and understanding tic-suppression-based behavior therapies for TS (10). We also speculate that tic timing patterns in patients with functional tics may be different than those seen in TS, potentially providing an objective tool to help with diagnosis (11, 12).

## Materials and Methods

Further details regarding the random walk model can be found in section S1 of the supporting information.

The clinical methods appear in detail elsewhere (13). Briefly, data from 78 children were used to generate 962 tic time series datapoints in this study. Children participated in the study within the first 6 months after onset of tics (screening visit) and at the 12 month anniversary of the first tic (12 month visit). At the 12 month visit, all children still had tics and most met diagnostic criteria for TS (14). At each visit, the timing of tics was recorded as described above. Author KJB recorded the timing of tics as they occurred, and rewards were delivered using a custom computer program connected to a token dispenser (15). In some participants, the NCR condition was omitted, so that most participants had 30-40 minutes of video at each visit.

All relevant datasets used for this study, including time series of tics can be found in supporting information datasets S1 and S2.

This study was approved by the Washington University Human Research Protection Office (IRB), protocol numbers 201109157 and 201707059. Each child assented and a parent (guardian) gave informed consent.

## Data Availability

All data and data processing codes are available upon request.

## Data Availability

Data is not generally available due to ethical considerations but may be requested from the authors.

## Acknowledgments

The authors would like to thank Drs. William R. Heinson, David Song, and Pai Liu.

