## Supporting Information for "Fractality of tics as a quantitative assessment tool for Tourette syndrome"

*Corresponding authors:

**This PDF file includes:**

Supplementary text

Legends for Datasets S1 to S2

**Other supplementary materials for this manuscript include the following:**

Datasets S1 to S2

Supplementary Information Text

**Directional Statistics for Random Walk Model.**

This work generates random walk trajectories according to directional statistics. The random walkers are first placed at position x = 0, and given an initial velocity of 1 position unit (arbitrary) per second. Time is then incremented by 0.1 seconds, and the walker will either reverse direction or move with the same velocity as the previous time step, according to a certain probability as a function of time (P_turn_(*t*)). The three schemes for random walk motion utilized in this work (ballistic motion, diffusive motion, and tic-modulated motion) are achieved by altering P_turn_(*t*).

Walkers undergoing so-called “tic-modulated” motion have P_turn_(*t*) which is determined by the tic time series. If a tic is not detected at a certain time *t*, then P_turn_(*t*) is equal to zero, and the walker continues moving with the same velocity as the previous time step. If a tic is detected at a certain time *t*, then P_turn_(*t*) is equal to unity, and the velocity of the walker is reversed. This is summarized by:

$$\text{P}_{\text{turn}}\text{(}\text{t}\text{)}\text{ }\text{=}\left\{ \begin{aligned} \text{ }\text{ }\text{0, no tic detected at }\text{t} \\ \text{1, tic detected at }\text{t} \end{aligned} \right.$$

Walkers undergoing diffusive motion have P_turn_(*t*) which is held constant at 0.5. This leads to a trajectory in which at any given time step, a walker is equally likely to continue moving with the same velocity as the previous time step, or have its velocity reversed. This type of motion is used to represent the worst-case scenario of a tic-modulated walk, where at any time step the patient is equally likely to tic or not tic. For diffusive motion, P_turn_(*t*) is given by:

$$\text{P}_{\text{turn}}\left( \text{t} \right)\text{ }\text{=}\text{ }\text{0.5}$$

Walkers undergoing ballistic motion have P_turn_(*t*) which is held constant at zero. This leads to a trajectory in which at any given time step, the walker continues moving with the same velocity as the previous time step. This type of motion is used to represent the best-case scenario of a tic-modulated walk, where at any time step the patient does not exhibit a tic. For ballistic motion, P_turn_(*t*) is given by:

$$\text{P}_{\text{turn}}\left( \text{t} \right)\text{ }\text{=}\text{ }\text{0}$$

Dataset S1 (separate .xlsx workbook) contains the following spreadsheets.

- Compiled Tic-Times: The time series of tics for the patients in this study.
  - Column 1. Header with format “patient_visit_session_suppression condition”
  - Subsequent columns show times where a tic was detected.
    - Units: seconds

Dataset S2 (separate .xlsx workbook) contains the following spreadsheets.

- Baseline (screening): Raw data for patients under baseline condition at screening visit.
  - Units:
    - Session, total tics, fractal dimension: unitless
    - Longest tic-free interval: seconds
- Baseline (12mo): Raw data for patients under baseline condition at 12 month follow-up visit.
  - Units:
    - Session, total tics, fractal dimension: unitless
    - Longest tic-free interval: seconds
- NCR (screening): Raw data for patients under NCR condition at screening visit.
  - Units:
    - Session, total tics, fractal dimension: unitless
    - Longest tic-free interval: seconds
- NCR (12mo): Raw data for patients under NCR condition at 12 month follow-up visit.
  - Units:
    - Session, total tics, fractal dimension: unitless
    - Longest tic-free interval: seconds
- Verbal (screening): Raw data for patients under verbal condition at screening visit.
  - Units:
    - Session, total tics, fractal dimension: unitless
    - Longest tic-free interval: seconds
- Verbal (12mo): Raw data for patients under verbal condition at 12 month follow-up visit.
  - Units:
    - Session, total tics, fractal dimension: unitless
    - Longest tic-free interval: seconds
- DRO (screening): Raw data for patients under DRO condition at screening visit.
  - Units:
    - Session, total tics, fractal dimension: unitless
    - Longest tic-free interval: seconds
- DRO (12mo): Raw data for patients under DRO condition at 12 month follow-up visit.
  - Units:
    - Session, total tics, fractal dimension: unitless
    - Longest tic-free interval: seconds
- TTS: Raw data for fractal dimension and corresponding total tic score (TTS) included in main text Figure 2a.
  - Units:
    - Fractal Dimension (screening visit): unitless
    - Fractal Dimension (12 month visit): unitless
    - TTS (screening visit): unitless
    - TTS (12 month visit): unitless
